# Liver biopsy confirms precise and efficient correction of SERPINA1 after *in vivo* Base Editing in a Patient with Alpha-1 Antitrypsin Deficiency

**DOI:** 10.64898/2026.06.01.26354551

**Authors:** Simon Alexander Krooss, Taihua Yang, Qinggong Yuan, Nora Drick, Malte Sgodda, Julia Held, Patrick Behrendt, Björn Hartleben, Rembert Koczulla, Xiaoying Ma, Ying Liu, Heiner Wedemeyer, Sabina Janciauskiene, Nataliya Di Donato, Tobias Cantz, ZiJun Wang, Yuxuan Wu, Marius Hoeper, Qiang Xia, Michael Ott

**Affiliations:** Department of Gastroenterology, Hepatology, Infectious Diseases and Endocrinology, Hannover Medical School, Hannover, Germany; PRACTIS Clinician Scientist Program, Dean’s Office for Academic Career Development, Hannover Medical School, Hannover, Germany; Department of Liver Surgery, Ren Ji Hospital, Shanghai Jiao Tong University School of Medicine, Shanghai, China; Sino-German Gene and cell therapy center, Ren Ji Hospital, Shanghai, China; Department of Respiratory Medicine and Infectious Diseases, Hannover Medical School, Hannover, Germany; Member of the German Center for Lung Research (DZL), Biomedical Research in Endstage and Obstructive Lung Disease Hanover (BREATH), Hannover, Germany; Institute for Experimental Virology, TWINCORE, Centre for Experimental and Clinical Infection Research, a Joint Venture Between the Helmholtz Centre for Infection Research and the Hannover Medical School, Hannover, Germany; DZIF, Partner Site Hannover-Braunschweig, Germany; Institute of Pathology, Hannover Medical School, Hannover, Germany; Institute for Pneumological Rehabilitation, Schön Klinik Berchtesgadener Land, Germany; YolTech Therapeutics, Shanghai, China; Department of Human Genetics, Hannover Medical School, Germany

**Keywords:** Alpha 1 Antitrypsin Deficiency, Adenine Base Editing, *in vivo* gene correction, gene therapy

## Abstract

**Background:** Alpha-1 antitrypsin deficiency (AATD) caused by the PI*ZZ mutation (Glu342Lys) results in hepatic accumulation of misfolded AAT-Z protein and reduced circulating AAT levels, leading to progressive liver disease and emphysema. Gene correction therapy represents a potentially curative approach by directly correcting the underlying genetic defect. We report the first case of successful hepatic gene correction with early histological and functional assessment.

**Methods/Case presentation:** We report the case of a 66-year-old male patient with PI*ZZ AATD who underwent gene correction therapy within the YOLT-202 phase I/Ia clinical trial (clinicaltrial.gov ID NCT07193615). Ten weeks post treatment a liver biopsy was performed to re-evaluate pre-existing F2 liver fibrosis as measured by elastography before entering the study. Serum samples allowed functional assessment of the AAT-mediated elastase inhibition.

**Results:** Liver biopsy did not show signs of hepatic inflammation and demonstrated 54% (Sanger) and 57% (Illumina) gene correction rate of the PI*ZZ variant on the DNA level with no bystander edits or off-target effects. Following a transient elevation of transaminases during the early post-treatment period, liver enzymes normalized. Monthly serum AAT measurements demonstrated biologically active and stable therapeutic levels throughout follow-up.

**Conclusions:** This case demonstrates efficient and precise hepatic gene correction without concerning histological alterations and with substantial improvement of functional parameters, supporting the feasibility and safety of gene editing approaches for AATD.

**One-sentence summary:** Liver biopsy after PI*ZZ gene correction therapy reveals efficient and accurate base editing.

## Introduction

Alpha-1 antitrypsin deficiency is caused by mutations in SERPINA1, with the PI*ZZ genotype accounting for about 95% of severe cases. The main clinical manifestations involve the lungs and the liver. Pulmonary disease typically presents as panacinar emphysema. Liver disease results from intrahepatic accumulation of the misfolded Z-AAT protein, which polymerizes within the endoplasmic reticulum of hepatocytes, forming periodic acid–Schiff with diastase (PAS-D)-positive inclusions that induce endoplasmic reticulum stress, hepatocellular injury, inflammation, and progressive hepatic fibrosis. The lifetime risk of cirrhosis in PI*ZZ individuals may reach 20-40%. [1]

Although gene therapy approaches using viral vectors have shown promise for pulmonary manifestations, achieving therapeutic AAT levels remains challenging.[2][3][4] Gene correction therapies, such as those using adenine base editors, offer the potential advantage of correcting the endogenous gene, thereby eliminating toxic Z-AAT production while restoring normal AAT synthesis. [5] Lipid nanoparticle (LNP)-delivered gene editing offers potential advantages including reduced immunogenicity, the possibility of repeat dosing, and transient expression of editing machinery that may improve the safety profile. Base editing technology enables precise single-nucleotide corrections without inducing double-strand breaks, potentially reducing the risk of off-target insertions or deletions. To date, no published reports have documented histological assessment of hepatic gene correction in AATD patients.

## Materials and Methods

### Liver Biopsy Procedure

A percutaneous liver biopsy was performed under ultrasound guidance using an 16G biopsy needle (Bard Access Systems, Covington, LA, USA). The biopsy was clinically indicated to further evaluate borderline elevated liver stiffness detected by noninvasive assessment and to confirm or exclude relevant hepatic fibrosis or cirrhosis. Written informed consent was obtained prior to the procedure. Following local infiltration with 10 mL Xylocaine 2%, a core specimen (approximately 15–20 mm in length) was obtained from the right hepatic lobe. The sample was immediately snap-frozen in liquid nitrogen and stored at −80 °C until further processing.

### Histological Staining and Analysis

Liver tissue sections (2 μm thick) were stained with periodic acid-Schiff after diastase treatment (D-PAS) to assess the presence of abnormal α1-antitrypsin (AAT) aggregates in hepatocytes. Stained slides were examined under light microscopy by a pathologist to evaluate morphological changes and the distribution of D-PAS-positive inclusions.

### Serum α1-Antitrypsin Quantification

Serum levels of AAT were measured using a cobas® photometric assay (Roche Diagnostics, Mannheim, Germany) on a cobas® 8000 analyzer (Roche Diagnostics). The assay was performed in accordance with the manufacturer’s instructions, and results were reported in µmol/l.

### Purification and Functional Analysis of Patient-Derived AAT

AAT was purified from patient plasma collected at multiple post-gene-therapy timepoints using affinity chromatography with an anti-AAT antibody-coupled Protein A/G resin. Following purification, AAT-containing fractions were dialyzed against phosphate-buffered saline (PBS, pH 7.4) and stored at −80 °C until further analysis.

Functional anti-elastase activity of purified AAT was assessed using a porcine pancreatic elastase (PPE) inhibition assay. Purified patient-derived AAT was preincubated with PPE for 5 min at 37 °C prior to initiation of the enzymatic reaction. Elastase activity was subsequently measured by monitoring substrate cleavage as the increase in optical density at 405 nm (OD_405_) every 8 s for 3 min using an Infinite® M200 microplate reader (Tecan). Anti-elastase activity was determined from the slope of the OD_405_ increase over time (ΔOD_405_/min), reflecting the rate of elastase-mediated substrate cleavage. Elastase activity in the absence of inhibitor (PPE alone) was defined as 100 % activity. Purified Z-AAT isolated from pooled plasma of seven Pi*ZZ patients and purified M-AAT (Respreeza®, CSL Behring, Marburg, Germany) were analyzed in parallel as controls under identical experimental conditions. All samples were measured in triplicate or quadruplicate (n = 4–6) to ensure reproducibility.

### DNA Extraction and Illumina Sequencing

Genomic DNA was isolated from the liver biopsy tissue using a VWR peq GOld Blood & Tissue DNA Mini Kit. DNA concentration was quantified using a Qubit®. Libraries for short read genome sequencing were prepared pcr-free xGen DNA Library Prep Kit EZ (IDT) on a Biomek FXp (Beckman Coulter), followed by 160 bp paired-end sequencing on an Illumina NovaSeq X Plus instrument. Insert size was in mean 314 bp, with a mean target region sequencing depth of 75x, generating 2,26 billion reads per sample. Raw sequencing data were processed using DRAGEN (Illumina) and megSAP platform. In addition, the predicted off-target regions as determined via GUIDE-seq and Digenome-seq (**Table 2**) were examined.

### Sanger Sequencing and BEAT Analysis

For targeted validation of base editing efficiency at the SERPINA1 locus, genomic DNA was extracted from the same biopsy sample using a commercial kit (Zymo Research #D7001). PCR amplification of the target region was performed using primers p1 (Fw: 5′-GACGTGGAGTGACGATGCT -3′) and p2 (Rv: 5′-TACCCAGCCAGATGCTCCAT -3′), flanking the edited site within exon 5 of SERPINA1. Amplification was carried out in a 50 μL reaction containing 1× PCR buffer Herculase2/using Herculase2 (Agilent #600677)

The PCR products were purified by gel electrophoreses (NEB Monarch DNA Gel Extraction Kit #T1020) sequenced using the same primers. Chromatograms were analyzed using the BEAT (Base Editing Analysis Tool) software (version 1.2), which enables quantitative assessment of base editing efficiency by deconvoluting mixed peaks at the target site. Editing efficiency was calculated as the percentage of corrected alleles (A→G transition at the target C) relative to the total number of reads. The results were compared to non-edited control sequences.

## Case Presentation

A 66-year-old male patient with genetically confirmed PI*ZZ AATD was evaluated for gene correction therapy. The diagnosis of AATD had been established 4 years before during diagnostic work-up for advanced pulmonary emphysema and subsequent SERPINA1 DNA sequencing confirming homozygosity for the Z allele (NM_000295.5:c.1096G>A p.Glu366Lys (HGVS)). Relevant comorbidities included arterial hypertension, well-controlled on antihypertensive therapy, and mild to moderate aortic valve insufficiency, which was hemodynamically stable and under regular echocardiographic surveillance. The patient had no history of tobacco use.

Comprehensive liver imaging was performed prior to treatment. Transient elastography (FibroScan) demonstrated a liver stiffness of 7.9 kPa, while acoustic radiation force impulse imaging (ARFI) revealed a shear wave velocity of 1.34 m/s, with both findings raising suspicion of underlying hepatic fibrosis.

Abdominal ultrasound revealed normal liver echotexture with undisturbed orthograde portal venous flow, excluding portal hypertension. No intra- or extrahepatic bile duct dilatation was observed. A known echogenic lesion in segment 7, previously characterized as a hemangioma by contrast-enhanced CT and ultrasound, remained stable in size and appearance as compared to years before.

### Gene Correction Therapy

On December 15, 2025, the patient received intravenous gene correction therapy with YOLT-202, a base editor formulated in lipid nanoparticles designed to correct the PI*ZZ mutation in SERPINA1 (ClinicalTrials.gov ID: NCT07193615). Premedication with oral dexamethasone were administered to mitigate potential immune responses to the lipid nanoparticle components. On the evening prior to treatment (December 14, 2025, at 20:00), the patient received oral dexamethasone. Within one hour before infusion on the treatment day, the patient additionally received intravenous dexamethasone sodium phosphate, oral compound acetaminophen tablets, oral levocetirizine hydrochloride tablets, and oral famotidine. The gene therapy product (YOLT-202, 35 mg) was diluted in 100 mL normal saline and administered as a single intravenous infusion over one hour. The infusion was well tolerated without immediate adverse events. The patient was monitored for three days post-infusion before discharge.

### Post treatment course

At two hours post-infusion, the patient has transiently developed subfebrile body temperature (maximum body temperature 37.2 °C). In addition, the patient developed transient transaminase elevation consistent with expected immune-mediated hepatotoxicity. Peak ALT of 60.9 U/L (1.35 × upper limit of normal [45 U/L]) and AST of 40.7 U/L (1.16 × ULN [35 U/L]) occurred at day 3 post-infusion (**Table 1**). Transaminases completely normalized at the third time point of blood withdrawal 14 days post infusion and remained normal during the remaining observation period.

**Table 1:** Serum biomarkers. **Longitudinal laboratory parameters before and after in vivo gene editing therapy**. Laboratory parameters were assessed 2 months before treatment, 3 days after treatment, 14 days after treatment, and at the time of liver biopsy. The liver biopsy was performed 10 weeks after administration of the investigational therapy. ALT, alanine aminotransferase; AST, aspartate aminotransferase; GGT, gamma-glutamyl transferase; CHE, cholinesterase.

|  | 2 months before treatment | 3 days post treatment | 14 days post treatment | 10 weeks post treatment |
| --- | --- | --- | --- | --- |
| **Serum biomarkers** |  |  |  |  |
| AAT [g/l] | 0.15 |  |  | 0.7 |
| CRP [mg/l] |  |  |  | <0.6 |
| AST [U/l] | 24 | 40.7 | 30.2 | 26 |
| ALT [U/l] | 23 | 60.9 | 32.6 | 22 |
| Alkaline Phosphatase [U/l] | 96 |  |  | 80 |
| GGT [U/l] | 15 |  |  | 11 |
| CHE [kU/l] | 6.93 |  |  | 7.16 |
| Bilirubin [μmol/l] | 8 | | | 13 |
| INR | 1.05 |  |  | 0.99 |

Continuous serum AAT measurements demonstrated stable levels above the therapeutic threshold of 0.57 g/l (**Figure 1A**). To verify the functionality of the patient’s enzyme, elastase inhibition capacity was measured. Strikingly, the elastase inhibition activity was comparable to the M-genotype control. (**Figure 1B)**. The histopathological analysis of the liver specimen revealed focal periportal D-PAS-positive hepatocellular inclusion bodies as a minor finding, along with mild signs of venous congestion with slight vascular and sinusoidal ectasia, minimal steatosis (5%), mild portal inflammation, and mild portal fibrosis (Ishak Scoring: A0, B0, C0, D1, F1). A representative section is depicted in **Figure 1C**. Subsequently, the residual tissue was objected to molecular analysis. Targeted amplicon sequencing confirmed A>G transition at the Z mutation site, restoring wild-type Glu342 codon. A quantification of the correction rate via BEAT analysis revealed 54% with no evidence for bystander-editing activity (**Figure 1C**). In addition, short-read genome sequencing was performed on the residual tissue demonstrating 57% corrected allele frequency and absence of bystander edits confirming the results obtained by Sanger sequencing (**Figure 1E**). Off-target sites, which were identified via GUIDE-seq and Digenome-seq in preclinical experiments, were assessed via high coverage genome and revealed no undesired edits (**Table 2**).

**Table 2:**
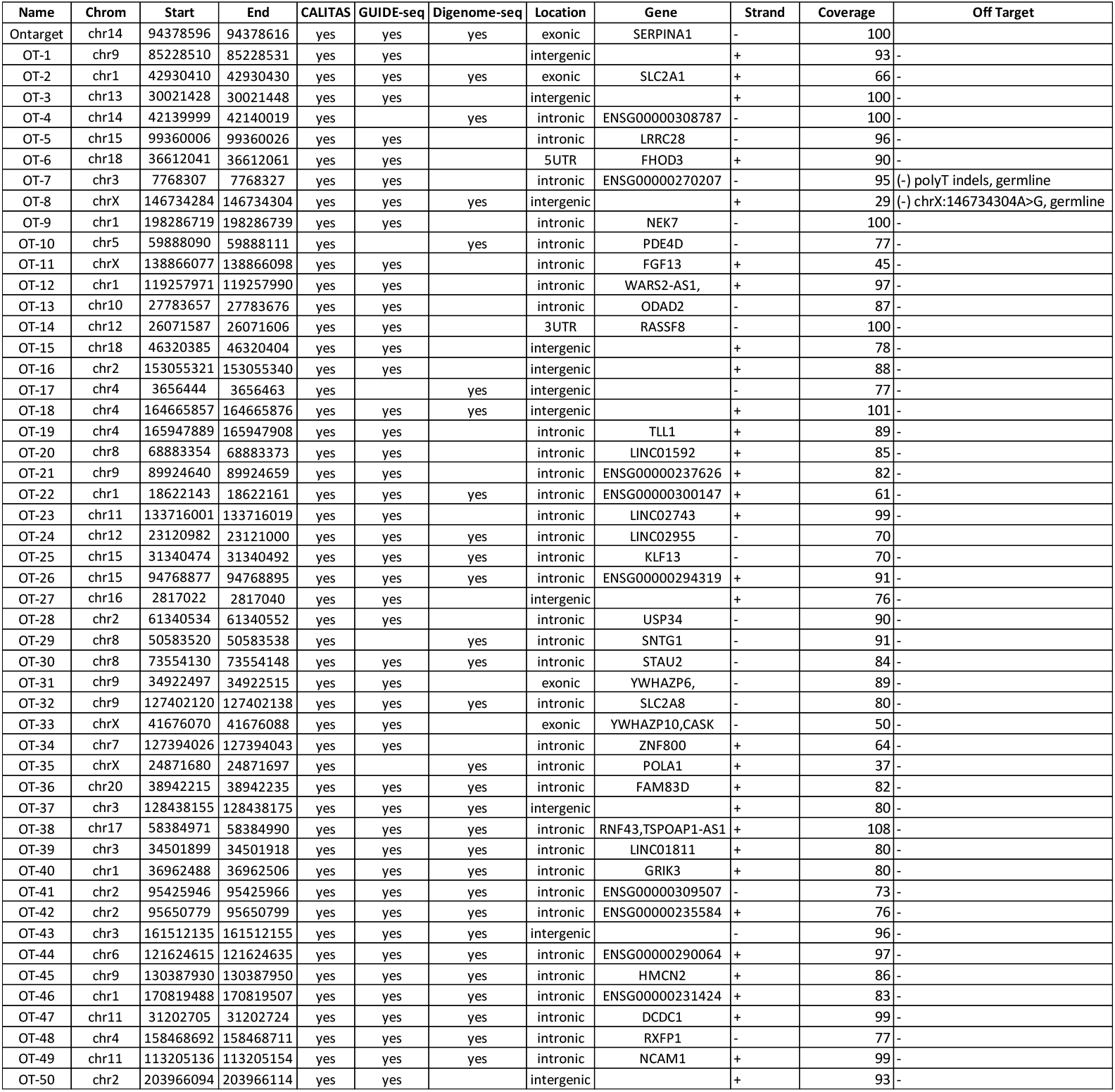
Analysis of predicted off-target editing sites in liver tissue following in vivo base editing therapy. Predicted off-target loci identified in preclinical studies by GUIDE-seq and Digenome-seq were analyzed in liver biopsy tissue obtained after treatment. Genomic DNA extracted from the biopsy specimen was subjected to targeted Illumina next-generation sequencing to assess editing outcomes at the corresponding genomic regions. Editing frequencies, including potential off-target and bystander edits, were determined by deep sequencing analysis. No detectable off-target modifications were observed at the analyzed loci.

**Figure 1.**
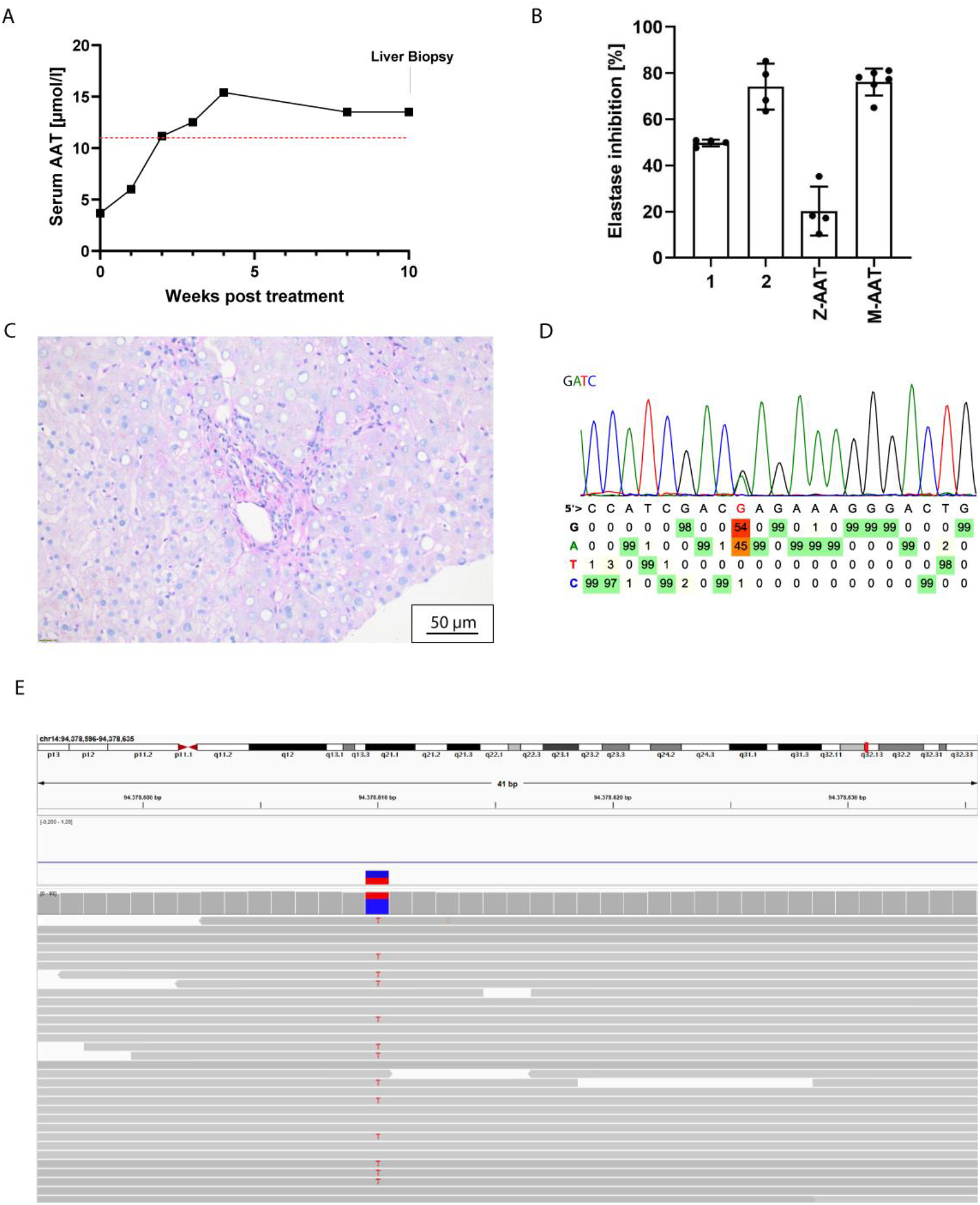
**(A)** Depiction of AAT levels in µmol/l after gene correction therapy. The red line indicates the protective threshold of 111 µmol/l. **(B)** Anti-elastase activity of affinity-purified patient AAT. Percentages of elastase inhibition were calculated from the slope of the absorbance increase at 405 nm over time (ΔOD_405_/min); the uninhibited reaction (elastase alone) was set to 100 %. Each sample was measured 4-6 times. **(C)** Representative liver section obtained 10 weeks post-treatment. High-magnification view confirms the near-absence of PAS-positive aggregates, with only minimal focal deposits observed. The histological pattern is consistent with a significant improvement in AAT clearance and hepatocellular function. The Ishak score for this biopsy is A0, B0, C0, D1, F1, reflecting minimal to no interface hepatitis (A0), no fibrosis (B0), no steatosis (C0), mild portal inflammation (D1), and mild ductular reaction (F1). The scale bar represents 50 µm. **(D)** Chromatogram and results of BEAT analysis indicating 54% correction efficiency and the absence of bystander edits in the surrounding A nucleotides. **(E)** IGV shown in reverse-complement reference orientation to match the SERPINA1 transcript strand; alignment reads remain displayed in genomic orientation. Variant corresponds to G>A on the transcript strand. Quantification of reads revealed 57% editing at the mutated position.

## Discussion

To our knowledge, this case demonstrates the first reported histological assessment of hepatic gene correction using a lipid nanoparticle-delivered base editor in a patient with PI*ZZ AATD. The 54-57% gene correction rate achieved at 2.5 months post-treatment with YOLT-202 demonstrates successful genomic editing in a substantial proportion of hepatocytes using a non-viral delivery platform. Given a correction rate of 57% of liver cell genomes, the number of corrected hepatocytes is expected to be even higher, indicating that the measured value likely represents a conservative estimate. Importantly, neither bystander edits nor off-target events were detected.

Based on the elevated FibroScan and ARFI measurements, both exceeding published cut-off values for liver fibrosis [6][7], a liver biopsy was performed to establish a definitive assessment of the underlying hepatic pathology.

The Ishak score of A0, B0, C0, D1, F1 indicates minimal disease activity with only mild portal inflammation and mild fibrosis. Importantly, there was no interface hepatitis (A0), no confluent necrosis (B0), and no focal lytic necrosis or apoptosis (C0), indicating absence of active hepatocellular injury. Mild portal inflammation (D1) with sparse lymphocytic infiltration and scattered ceroid-laden macrophages likely represent residual changes from prior disease activity rather than ongoing inflammation.

The histological findings at 2.5 months show no evidence of immune-mediated hepatitis related to gene therapy. Despite the transient transaminase elevation observed in the early post-treatment period, the liver biopsy demonstrates resolution of any acute inflammatory response. The achievement and maintenance of serum AAT levels above the protective threshold of 11 μmol/L (0.57 g/L) throughout the observation period is clinically significant. While the prognostic value of this threshold for risk stratification remains controversial, it is well established as a therapeutic target in AAT augmentation therapy.[8] The stability of AAT levels across monthly measurements suggests durable transgene expression from corrected hepatocytes and indicates a functional reversion of towards a PI*MZ genotype.

Taken together, the lack of detectable off-target effects and functionally relevant bystander edits across all analyses performed highlights the high precision of this approach and supports its favorable safety profile.

## Data Availability

All data produced in the present study are available upon reasonable request to the authors.

## Informed Consent

Human material and data used in this study was obtained in accordance with the principles of the Declaration of Helsinki, has been approved by the ethics committee of Hannover Medical School (No 2967-2015) and the patient provided written informed consent.

## Author contributions

S.A.K. conceived the idea, and wrote the manuscript; Y.T. and Q.X. supervised the clinical trial, X.M., Y.L., Z.W. and Y.W. coordinated the clinical trial; S.A.K., N.D., Q.Y., R.K., M.H. and M.O. coordinated patient acquisition and follow up; N.D.D. performed NGS-analysis; M.S., J.H., S.J. performed experiments, B.H. performed histological analysis. T.C., H.W., N.D., M.H., M.O., N.D.D. and T.Y. revised the manuscript.

## Acknowledgements

We thank the patient for providing permission to publish our findings.

## Supplemental Information

**Figure S1.**
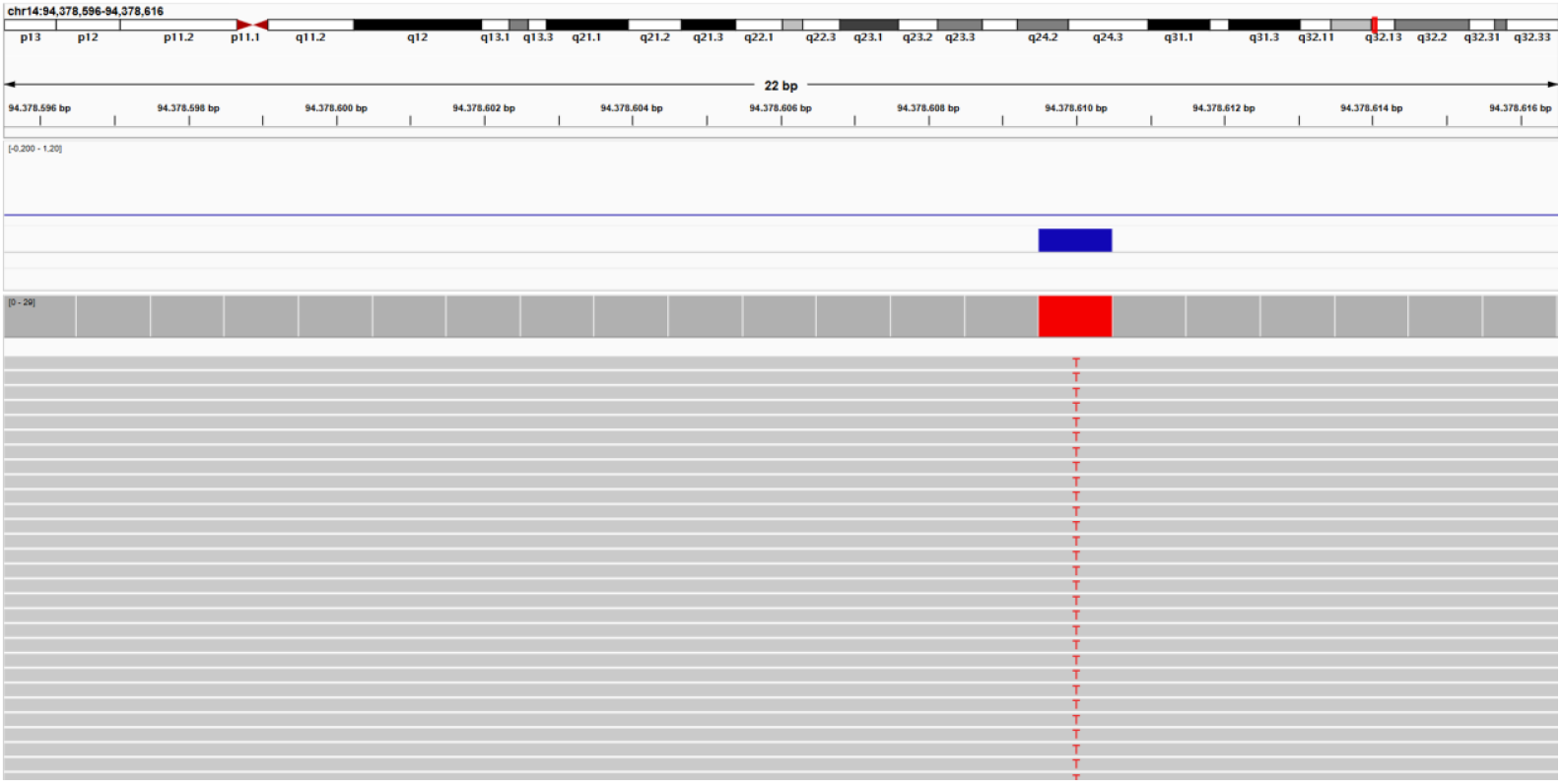
Depiction of sequencing data obtained from DNA of peripheral blood cells. This data served as a reference sequence for the analysis of the sequencing data obtained from the liver tissue. IGV shown in reverse complement reference indicating 100% presence of the PI*ZZ mutation.

